# Characterization of heart rate changes associated with autonomic dysreflexia during penile vibrostimulation and urodynamics

**DOI:** 10.1101/2021.04.03.21254489

**Authors:** Lauren Rietchel, Andrea L. Ramirez, Shea Hocaloski, Stacy Elliott, Matthias Walter, Andrei V. Krassioukov

## Abstract

**Purpose:** Autonomic dysreflexia, often accompanied by heart rate changes, increases the risk of cardio-cerebrovascular complications in individuals with spinal cord injury. Thus, our aim was to characterize these changes during penile vibrostimulation and urodynamics.

**Materials and Methods:** We analyzed the cardiovascular (i.e. blood pressure and heart rate) data from two prospective studies, i.e. 21 individuals with chronic spinal cord injuries and history of autonomic dysreflexia, who underwent penile vibrostimulation (n=11, study 1) or urodynamics (n=10, study 2).

**Results:** The cohort’s median age was 41 years (range 22 −53). Overall 47 episodes of autonomic dysreflexia were recorded (i.e. penile vibrostimulation n=37, urodynamics n=10), while at least one episode was recorded in each participant. At the threshold of autonomic dysreflexia, bradycardia was observed during penile vibrostimulation and urodynamics in 43% and 30% of all episodes, respectively. At the peak of autonomic dysreflexia during penile vibrostimulation and urodynamics, bradycardia was observed in 65% and 50%, respectively. In contrast, tachycardia was detected only once during urodynamics.

**Conclusion:** Our findings reveal that heart rate changes associated with autonomic dysreflexia during penile vibrostimulation and urodynamics appear to be related to the magnitude of systolic blood pressure increases. Thus, highly elevated systolic blood pressure associated with bradycardia suggest the presence of severe autonomic dysreflexia, which can lead to devastating cerebro-cardiovascular consequences. Therefore, we recommend cardiovascular monitoring during penile vibrostimulation and urodynamics to detect autonomic dysreflexia and stop assessments before systolic blood pressure is dangerously increasing, thereby reducing the risk of potentially life-threatening complications in this cohort.

## INTRODUCTION

Long-term consequences of SCI are devastating and extensive.^1^ Of note, autonomic functions, including cardiovascular, LUT and sexual function are often impaired post-injury, thereby placing a tremendous burden on this population.^2,3^ AD is a well-known autonomic consequence following SCI^4^, which, if not treated, can lead to life-threatening complications including stroke, myocardial infarction or death.^5^

In daily life, AD can occur in response to intrinsic stimuli (e.g., bladder distention and UTI) and/or extrinsic stimuli (e.g., restrictive clothing).^6^ Although highly informative, commonly performed urological diagnostic procedures (e.g., PVS, UDS, and cystoscopy) can trigger severe AD episodes^67,8^ and associated cardiac arrhythmias.^9^ PVS is a well-established technique for sperm retrieval in males who wish to father children.^10,11^ However, due to its intense stimulation, PVS is associated with high cardiovascular risk.^12^ UDS, the gold standard to assess ANLUTD in SCI, is also well-known to elicit AD.^13,14^

Currently, HR responses associated with AD during both procedures are indeterminate. Although most studies reported HR decreases (i.e., relative bradycardia, i.e. <60bpm,^5,8,9,15–17^) as the primary response during AD, which is in line with our understanding of the pathophysiologic HR and BP changes^18,19^ (i.e., a parasympathetically-mediated HR decrease in order to lower SBP during AD ^2,7,9,20–22^), Solinsky et al. reported that relative tachycardia (i.e., >100bpm), not bradycardia (i.e. 68% vs. 0.3%) was the primary HR response during AD.^23^ Given this conflicting literature, we aimed to characterize iatrogenically induced AD (i.e. during urodynamics or sperm retrieval) associated HR changes during both procedures using continuous beat-to-beat BP monitoring, a monitoring system crucial in order to capture peak systolic change.^12^ Considering the lack of official safety protocols and the evidence of well-known cardio-cerebrovascular complications associated with AD, better knowledge of cardiovascular changes associated with AD (e.g., HR changes) is essential to improve the safety of individuals with SCI.

## MATERIALS AND METHODS

### Study design and ethics

This is an analysis of complete cardiovascular data sets from two prospective studies (i.e., one study not qualifying as an applicable clinical trial, and one clinical trial – NCT02676154), approved by the University of British Columbia Clinical Research Ethics Board, Vancouver Coastal Health Research Institute and Health Canada.

### Participants

We screened for complete records of individuals with chronic SCI and NLI at or above the sixth thoracic spinal level who underwent PVS or UDS. Complete inclusion and exclusion criteria, study design and diagnostic assessments are described in the supplementary methods.

### Outcome

Outcome was the characterization of HR changes (i.e. increase, decrease, tachycardia or bradycardia) at AD threshold (i.e., change [Δ] in SBP >20mmHg from baseline) and AD peak (i.e. maximum ΔSBP during AD). In accordance with the ISAFSCI, AD was defined as an increase in SBP of >20 mmHg from baseline.^6^

### Hemodynamic Assessments

Cardiovascular parameters were recorded continuously for ten minutes before (i.e., baseline) and during the UDS and PVS. HR (ECG, lead II; Powerlab Model ML132, ADInstruments, Colorado Springs, Colorado) and beat-to-beat BP (via photoplethysmography; Finometer, Finapres Medical Systems BV, Arnhem, The Netherlands) were monitored non-invasively. Data were collected at a rate of 1kHz per channel through an analog to digital converter (Powerlab/16SBP model ML795, ADInstruments, Colorado Springs, Colorado) and calibrated to brachial BP measurements (Dinamap 100, GE, Fairfield, Connecticut, USA) taken in 1-minute intervals throughout each procedure. The time to and the duration of AD were recorded during PVS. Continuous data collection occurred throughout each procedure using LabChart^®^ software (Version 8.0, ADInstruments, Colorado Springs, Colorado). All figures were created with Prism Version 8.4.0 (GraphPad Software, San Diego, California). The protocols, devices used, and data extraction, are explained in the supplementary methods.

### Statistical Analysis

Statistical analyses were conducted using R Statistical Software Version 3.6.0 for Macintosh Operating System. Data are presented as raw values and percentage, and median with IQR. Range (i.e., min -max) is provided for age, time post-injury, and number of PVS per individual. In LabChart^®^ software, raw data were extracted as a 3-beat average per point based on the ECG. From these averaged data points, the cardiovascular parameters from one minute prior to recording were calculated as the average (i.e., mean). For each AD episode during PVS and UDS, AD threshold and AD peak were identified. Then, all corresponding HRs were identified to calculate ΔHR from baseline, i.e., at AD threshold and AD peak. In PVS only, baseline parameters were compared to the relative response for each stimulation, and new baseline data were established for each stimulation. AD threshold for each stimulation was identified based on each re-calculated baseline. The median and IQR were calculated individually for each PVS, and all stimulations taken together. SBP and HR were also determined at the point of ejaculation for eligible participants. In addition, the median and IQR of the grouped data are reported. In UDS only, we analyzed documented clinical signs and symptoms.

## RESULTS

### Participants

In total, 21 individuals (median age 41 years [IQR 37–47, range 22–53], with a median post-injury time of 18 years [IQR 7–27, range 4–39]), who underwent PVS (11/21, all male) or UDS (10/21, including 4 females) with complete recordings were included in our analysis. Demographics and injury characteristics of participants are shown in table 1.

**Table 1:**
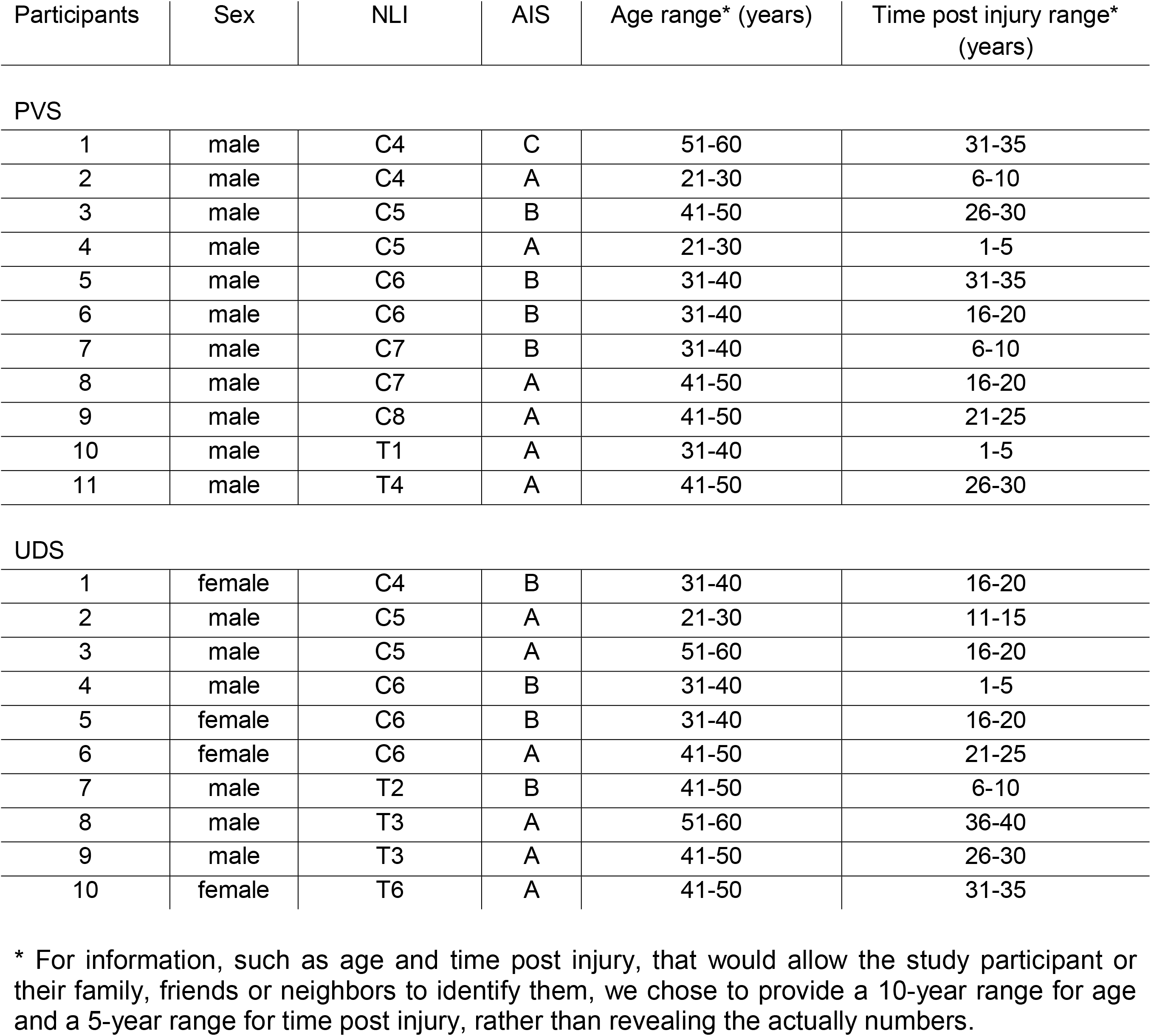
Participant’s demographics and injury characteristics.

| Participants | Sex | NLI | AIS | Age range* (years) | Time post injury range* (years) |
| --- | --- | --- | --- | --- | --- |
| PVS |  |  |  |  |  |
| 1 | male | C4 | C | 51-60 | 31-35 |
| 2 | male | C4 | A | 21-30 | 6-10 |
| 3 | male | C5 | B | 41-50 | 26-30 |
| 4 | male | C5 | A | 21-30 | 1-5 |
| 5 | male | C6 | B | 31-40 | 31-35 |
| 6 | male | C6 | B | 31-40 | 16-20 |
| 7 | male | C7 | B | 31-40 | 6-10 |
| 8 | male | C7 | A | 41-50 | 16-20 |
| 9 | male | C8 | A | 41-50 | 21-25 |
| 10 | male | T1 | A | 31-40 | 1-5 |
| 11 | male | T4 | A | 41-50 | 26-30 |
| UDS |  |  |  |  |  |
| 1 | female | C4 | B | 31-40 | 16-20 |
| 2 | male | C5 | A | 21-30 | 11-15 |
| 3 | male | C5 | A | 51-60 | 16-20 |
| 4 | male | C6 | B | 31-40 | 1-5 |
| 5 | female | C6 | B | 31-40 | 16-20 |
| 6 | female | C6 | A | 41-50 | 21-25 |
| 7 | male | T2 | B | 41-50 | 6-10 |
| 8 | male | T3 | A | 51-60 | 36-40 |
| 9 | male | T3 | A | 41-50 | 26-30 |
| 10 | female | T6 | A | 41-50 | 31-35 |

\* For information, such as age and time post injury, that would allow the study participant or their family, friends or neighbors to identify them, we chose to provide a 10-year range for age and a 5-year range for time post injury, rather than revealing the actually numbers.

### Cardiovascular parameters

Table 2 provides an overview of SBP and HR values at baseline, at AD threshold and at AD peak including their changes where appropriate.

**Table 2:** Overview of cardiovascular parameter and their changes during PVS and UDS

| | Baseline | | At AD threshold | | | At AD peak (i.e. maximum ΔSBP from baseline) | | | |
| --- | --- | --- | --- | --- | --- | --- | --- | --- | --- |
| | SBP [mmHg] | HR [bpm] | SBP [mmHg] | HR [bpm] | ΔHR [bpm] | SBP [mmHg] | ΔSBP [mmHg] | HR [bpm] | ΔHR [bpm] |
| **PVS** |  |  |  |  |  |  |  |  |  |
| 1 (n=11) | 113 [97 – 121] | 70 [62 – 76] | 139 [125 – 144] | 64 [59 – 76] | 0 [-4 – 5] | 170 [164 – 199] | 65 [51 – 85] | 57 [45 – 71] | -17 [-19 – -4] |
| 2 (n=10) | 113 [103 – 122] | 64 [56 – 70] | 143 [129 – 147] | 60 [55 – 69] | 0 [-1 – 4] | 190 [180 – 206] | 67 [55 – 95] | 51 [47 – 62] | -8 [-14 – -6] |
| 3 (n=9) | 111 [110 – 124] | 62 [56 – 72] | 143 [135 – 158] | 57 [54 – 67] | 1 [-5 – 5] | 196 [175 – 218] | 70 [51 – 107] | 53 [52 – 68] | -9 [-16 – -4] |
| 4 (n=6) | 119 [114 – 120] | 61 [60 – 70] | 144 [138 – 148] | 64 [56 – 72] | 1 [-5 – 2] | 192 [156 – 227] | 65 [31 – 113] | 56 [54 – 66] | -9 [-14 – -6] |
| 5 (n=1) | 118 | 70 | 143 | 86 | 17 | 246 | 128 | 86 | -21 |
| **UDS** |  |  |  |  |  |  |  |  |  |
| 1 (n=10) | 120 [112 – 128] | 73 [59 – 74] | 145 [133 – 155] | 65 [55 – 83] | -1 [-5 – 2] | 179 [169 – 185] | 53 [43 – 75] | 60 [52 – 73] | -2 [-13 – -1] |

Per assessment an individual underwent a maximum of five PVS or one UDS. All values are represented as the median and associated quartiles. The HR from baseline to ‘at AD threshold’ is reflective of the median for each, and the change in heart rate reflects the median of all of the HR changes, so although the median HR changed from a “Baseline” of 70bpm to 64bpm “at AD threshold”, the median change for all individuals combined is 0 as that was the median value. It is not to say that individual HR’s did not increase and decrease compared to baseline at AD threshold.

#### Penile vibrostimulation

During an average of 3 PVS per participant (range 1 to 5), we identified 37 AD episodes across all individuals. Figure 1 depicts the individual ΔSBP during each PVS (data shown in Table 2) as well as HR changes at AD threshold and peak AD. At baseline, median SBP and HR were 114mmHg (104 – 120) and 65bpm (59 – 73) for all 37 AD episodes. Upon reaching AD threshold, the median HR was 62bpm (56 – 76) with a median ΔHR of 1bpm (−5 – 5). At AD threshold, HR increased in 51% (19/37, median Δ = 5bpm, 2 – 8), decreased in 41% (15/37, median Δ = −7bpm, −13 – −3) or remained unchanged in 8% (3/37). Furthermore, bradycardia was present in 43% episodes (16/37, median HR 55bpm, 52 – 57), while in 57% (21/37) median HR was 76bpm (64 – 80). At AD peak (median ΔSBP 67mmHg, 51 – 99), an associated median HR (−10bpm, −17 – −5) was recorded across all participants. As well, the majority of HR changes were classified as bradycardia (65%, 24/37, median 51bpm, 43-54), while the remaining 35% (13/37) HR changes remained below the threshold for tachycardia (median 72bpm, 69 – 73). The moment of ejaculation during PVS was captured in 4 individuals, revealing a median ΔSBP of 98mmHg (84 – 123) at AD peak with an associated median ΔHR of −19bpm (−23 – −17).

**Figure 1.**
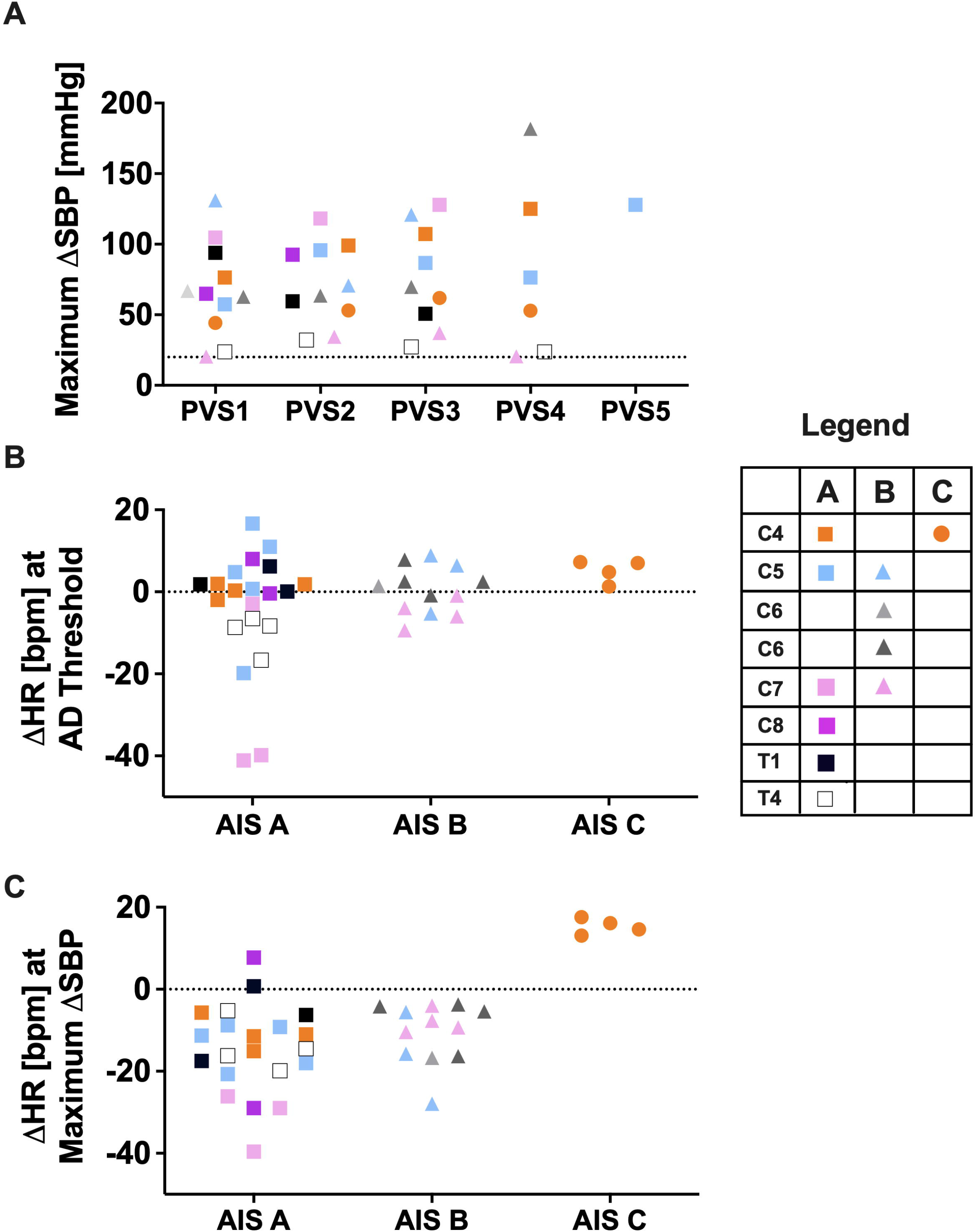
Cardiovascular changes during PVS. A) Depicts the individual’s maximum increase (Δ) in SBP during each PVS. The dotted line represents AD threshold (i.e. SBP ≥ 20mmHg). B) Represents the participant’s change in HR at the AD threshold across all PVS distributed according to the severity of SCI (i.e. AIS). At AD threshold, HR increased in 51% (19/37, median Δ = 5bpm, 2 – 8), decreased in 41% (15/37, median Δ = −7bpm, −13 – −3) or remained unchanged in 8% (3/37). The dotted line represents the HR at baseline. C) Shows the participant’s change in HR at AD peak (i.e. the maximum (Δ) SBP) across all PVS distributed according to the severity of SCI. At AD peak, 84% (31/37) of AD episodes, median ΔSBP of 76mmHg (47–106), were associated with a decrease in median HR of −12bpm (−18– −7) for the 10 motor-complete (AIS A/B) individuals. Among those, two individuals each had an individual PVS episode which showed an increase in ΔHR at the peak of AD (2/37; ΔHR = 1bpm for a T1 AIS A participant, and ΔHR = 8bpm for a C8 AIS A participant), although the remainder of their respective PVS AD episodes showed a decrease in ΔHR at AD peak. The only one participant with a C4, motor-incomplete (i.e. AIS C) injury (depicted as an orange circle) consistently showed increased HR [median Δ15bpm (14–17)] at AD peak across the 4 PVS. The dotted line represents the HR at baseline. The participants legend highlights the level and severity of their SCI.

#### Urodynamics

During a total of 10 UDS, we identified an episode of AD in each individual (all motor-complete SCI, i.e. AIS A/B). Figure 2 depicts the individual ΔSBP during each UDS (data shown in Table 2) as well as HR changes at AD threshold and peak. At AD threshold, a median HR of 65bpm (55 – 83) and median ΔHR of 0bpm (−3 – 2) was see across all participants. Furthermore, HR increased in 40% (4/10, median Δ = 3bpm, 3 – 7), decreased in 40% (4/10, median Δ = −4bpm, −6 – −3) or remained unchanged in 20% (2/10). Bradycardia was observed for 30% of episodes (3/10, median HR = 53bpm, 52 – 53), and the remaining 70% (7/10) of episodes showed a median HR of 79bpm (71 – 81). At AD peak (median ΔSBP 53mmHg, 43–75), an associated median HR (−1bpm, −8 – 1) was recorded. In 7 individuals an AD-associated HR decrease was observed. Five individuals (median = 53bpm, IQR = 48-55) experienced bradycardia, while 4/10 had a median HR of 73bpm (66 – 75). Only one individual (T3 AIS A), experienced tachycardia (102bpm) at AD peak. Table 3 summarizes the signs and symptoms of AD during the UDS.

**Table 3.** Clinical signs and symptoms experienced during UDS

| **Clinical signs and symptoms of AD during UDS** | **Number of participants** |
| --- | --- |
| *Sweating*<sup>\$</sup> | 3 |
| *Flushing*<sup>\$</sup> | 1 |
| *Headache*<sup>#</sup> | 1 |
| *A “rush to the head”* | 1 |
| *Tingling* | 1 |
| **Non-classic AD signs and symptoms noted during UDS** | **Number of participants** |
| *Unspecified bladder sensations* | 4 |
| *Unspecified spasms* | 3 |
| *Abdominal pressure* | 2 |
| *A sore lower back* | 1 |

<sup>\$</sup> In individuals (4/10) experiencing flushing and/or sweating, symptoms were identified at a median ΔSBP of 24mmHg (22 – 25) with a median ΔHR of 1bpm.
<sup>#</sup> One subject, who reported headache during UDS at a maximum ΔSBP of 31mmHg had an AD-associated median HR decrease of -2bpm.

**Figure 2.**
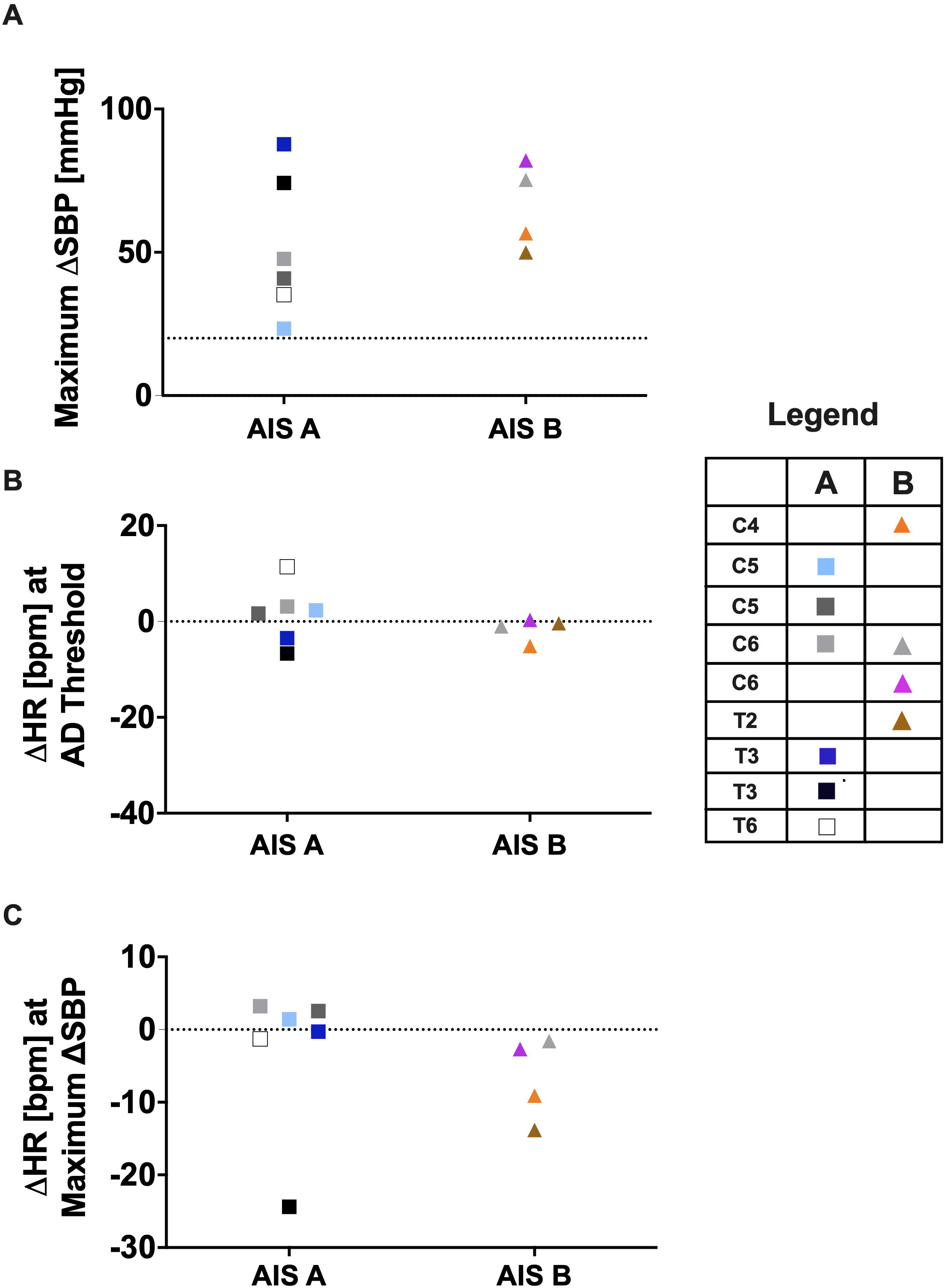
Cardiovascular changes during UDS. A) Depicts the individual’s maximum increase (Δ) in SBP during each UDS (n=10). The dotted line represents AD threshold (i.e. SBP ≥ 20mmHg). B) Represents the participant’s change in HR at the AD threshold across all UDS distributed according to the severity of SCI (i.e. AIS). The dotted line represents the HR at baseline. C) Shows the participant’s change in HR at the maximum (Δ) SBP across all UDS distributed according to the severity of SCI. The dotted line represents the HR at baseline. The participants legend highlights the level and severity of their SCI.

## DISCUSSION

### Main findings

The primary HR response at AD peak (i.e., compared to baseline) during PVS and UDS was a decrease in HR. Bradycardia, not tachycardia was mainly observed during PVS and in half the time during UDS. Only one participant had a tachycardia at AD peak during UDS. This pattern of AD associated HR change during both assessments appears to be dependent on the ΔSBP.

### Findings in context of existing evidence

The current findings are in line with the previous understanding of a parasympathetically-mediated response to counterbalance extensive SBP increases during AD.^15^ Considering the impairment of bulbospinal pathways following SCI, inhibitory efferent signals cannot reach and neutralize the activated SPNs below the NLI.^24^ Thus, vasoconstriction and thereby increased SBP are maintained as long as the AD stimulus is activated. Consequently, baroreceptors are activated, resulting in a decrease of HR, which can result in bradycardia and even asystole.^25,26^ Several research groups have observed and reported this pattern.^8,9,15,16,22^ Our observation of HR decreases at the peak of AD displays an impaired supraspinal control of the heart and SPNs below the NLI, accompanied by unopposed parasympathetical (i.e. vagal) tone during AD.^9^ In this current study, SBP reached as high as 300mmHg during PVS, potentially revealing the SBP-dependent effect on HR that was not shown in the 2018 study.^23^

The current study purposefully utilized continuous beat-to-beat BP monitoring to ensure greater accuracy of SBP peaks during AD episodes^12^, compared to intermittent BP monitoring. Continuous BP monitoring increases the validity of our findings by accurately detecting rapid and extreme SBP fluctuations.

Additionally, these HR findings are contrary to those from Solinsky et al., who reported relative tachycardia as the primary HR response during AD.^23^ This 2018 study was different from the design of our current study in that it had a more conservative definition of AD and no specific interventions. Our study measured SBP specifically during highly stimulating interventions. Furthermore, the Solinsky study measured BP manually and intermittently versus continuously, reducing the accuracy of the result and possibly underestimating the true SBP value.

However, this secondary analysis was neither powered nor designed to investigate HR changes specific to the NLI or severity (i.e. AIS grading). Nevertheless, we showed in a previous study that not only the higher the NLI, the greater the odds are to experience AD during UDS but those with a complete (AIS A) vs. incomplete SCI (AIS B – D) had greater AD-associated HR decreases.^7^

When considering the completeness of injury (i.e., AIS grade) and its impact on ΔHR and ΔSBP (i.e., AD severity), it is suggested that incomplete injuries will show better preserved cardiovascular function through reduced incidence of AD and increased low-frequency and high-frequency HR variability at rest.^27^ In both, UDS and PVS analyses there was high variability in ΔSBP and ΔHR response when considering completeness and level of injury. Our studies were not aimed at investigating HR changes specific to completeness of NLI, but instead to provide first insights to be thoroughly investigated in a future prospective trial.

In the UDS study only, AD symptoms were documented in all participants. This was done specifically in support of the findings of a 2017 study, showing that a patients age (>45 years), completeness of injury, and the symptoms of chills or sweating, are risk factors for AD symptoms occurring during UDS.^28^ In our study all participants had a complete SCI (i.e., AIS A or B), half the cohort was over 45 years of age, and 30% presented with sweating during UDS. Risk factor prediction of AD symptoms is a potentially useful tool in the clinical setting especially as patients are able to state their symptoms throughout a procedure. Additionally, a 2018 study suggested that neurogenic detrusor overactivity was also a predictor for AD occurring during UDS.^7^ The current UDS study supports these findings, where all participants exhibited neurogenic detrusor overactivity and AD during the procedure.

### Implications

These humble, novel findings might help inform the management of individuals with AD undergoing PVS and UDS, potentially other diagnostic procedures that could provoke AD. Although beat-to-beat cardiovascular monitoring is considered ideal, these devices are mainly found in research institutes. However, monitoring SBP every minute, as a minimal requirement for safety surveillance with regard to AD, seems feasible in a busy daily clinic. Since AD can be severe and asymptomatic in individuals undergoing PVS^22,29^ and UDS^7^, these findings may also aid to an increased awareness that rapid SBP increases can occur without clinical signs and symptoms, and without SBP monitoring would remain undetected and potentially lead to devastating cardiovascular consequences. Additionally, monitoring HR along with SBP during these procedures may help to recognize a profound and potentially dangerous episode of AD and thereby assist in knowing when to implement a safe stop protocol. Clinical signs and symptoms, including patient age, NLI and personal characteristics for prediction of AD can be utilized in a similar way to encourage a safer outcome.

Future research should focus on elucidating the mechanisms behind these HR responses, in addition to obtaining a wider range of data from different NLI or AIS grade. Future studies applying this knowledge in a clinical environment will be helpful for addressing its practical applications.

## Study limitations

Our study was limited by a small cohort of participants and the distribution of sex and injury characteristics. The small sample size limits the conclusion drawn from the NLI and its effect on HR outcomes in AD. The majority of participants were males (81%, 17/21), limiting the findings of our female cohort. Except for one participant with a motor -incomplete SCI (i.e. AIS C), all participants had a motor -complete SCI (AIS A / B), limiting the application of our findings solely to the latter group. Furthermore, our study participants all had chronic injuries, so we cannot comment on the characteristics of HR responses during PVS and UDS in patients with acute SCI. Additionally, there were four methods used during the PVS procedures, each with different modes and intensities, with the goal of maximizing the occurrence of ejaculation, which may have influenced the degree of AD experienced by participants in the PVS study. Given these limitations, further studies that include a greater variety of levels and completeness of injury would help to inform and individualize the HR response in each type of injury.

## CONCLUSION

This secondary analysis was aimed at characterizing the HR responses throughout AD to the point of maximum SBP increase (i.e. peak AD) in two common diagnostic procedures applied in daily urological practice potentially eliciting an AD episode. The findings document clinical bradycardia during AD as the primary response during the peak of AD in individuals with a motor-complete SCI, creating new implications for the management of AD during PVS and UDS. Although cardiovascular monitoring during these assessments is already recommended to reduce the risk of life-threatening complications of AD^5^, there is a potentially novel role for ΔHR to help characterize and identify peak AD. Considering that AD severity during UDS and PVS is supported by a decrease in HR, which seems to culminate at maximum ΔSBP, monitoring HR along with SBP may help identify the development of a serious AD episode in this cohort.

## Data Availability

The data are available from the corresponding author on reasonable request.

## ABBREVIATIONS

AD: autonomic dysreflexia
AIS: American Spinal Injury Association Impairment Scale
ANLUTD: adult neurogenic lower urinary tract dysfunction
BP: blood pressure
ECG: electrocardiogram
HR: heart rate
IQR: interquartile range
ISAFSCI: International standards to document remaining autonomic function after Spinal cord injury
LUT: lower urinary tract
NLI: neurological level of injury
PVS: penile vibrostimulation
QOL: quality of life
SBP: systolic blood pressure
SCI: spinal cord injury
SPN: sympathetic preganglionic neuron
UDS: urodynamic studies
UTI: urinary tract infection

## Acknowledgements

We wish to acknowledge the Blusson Spinal Cord Centre. Furthermore, we would like to thank the participants who provided written informed consent for participation and publication in this study.

## Author contributions

Andrei V. Krassioukov and Matthias Walter had full access to all the data in the study and take responsibility for the integrity of the data and the accuracy of the data analysis.

## Study concept and design

Lauren Rietchel, Andrea L. Ramirez, Stacy Elliott, Andrei V. Krassioukov, Matthias Walter

### Acquisition of data

Andrea L. Ramirez, Shea Hocaloski, Stacy Elliott, Andrei V. Krassioukov, Matthias Walter

### Analysis and interpretation of data

Lauren Rietchel, Andrea L. Ramirez, Shea Hocaloski, Stacy Elliott, Andrei V. Krassioukov, Matthias Walter

### Drafting of the manuscript

Lauren Rietchel

### Critical revision of the manuscript for important intellectual content

Lauren Rietchel, Andrea L. Ramirez, Shea Hocaloski, Stacy Elliott, Andrei V. Krassioukov, Matthias Walter

### Statistical analysis

Lauren Rietchel and Matthias Walter

### Supervision

Andrei V. Krassioukov and Matthias Walter

## Author responsibilities

According to the ICJME authorship, all authors provided substantial contributions to the conception or design of the work, or the acquisition, analysis or interpretation of data for the work; **AND** drafting the work or revising it critically for important intellectual content; **AND** final approval of the version to be published; **AND** agreement to be accountable for all aspects of the work in ensuring that questions related to the accuracy or integrity of any part of the work are appropriately investigated and resolved.

## Obtaining funding

This study was funded by the Rick Hansen Man in Motion Research Foundation (grant number: 135774, awarded to Dr. Krassioukov). The research equipment for this study was supported by the Canadian Foundation of Innovation (CFI, grant number: 35869) and the British Columbia Knowledge Development Fund (BCKDF, grant number: 35869. Dr. Walter was supported by a 2017-2019 Michael Smith Foundation for Health Research (MSFHR) and Rick Hansen Foundation Postdoctoral Research Trainee Award (grant number 17110). Dr. Krassioukov holds the Endowed Chair in Rehabilitation Medicine.

## REFERENCES

1. McKinley WO, Jackson AB, Cardenas DD, DeVivo MJ. Long-term medical complications after traumatic spinal cord injury: A Regional Model Systems Analysis. Arch Phys Med Rehabil. 1999;80(11):1402–1410. doi:10.1016/S0003-9993(99)90251-4

2. Lindan R, Joiner E, Freehafer AA, Hazel C. Incidence and clinical features of autonomic dysreflexia in patients with spinal cord injury. Paraplegia. 1980;18(5):285–292. doi:10.1038/sc.1980.51

3. Karlsson AK. Autonomic dysreflexia. Spinal Cord. 1999;37(6):383–391.

4. Lee ES, Joo MC. Prevalence of Autonomic Dysreflexia in Patients with Spinal Cord Injury above T6. Biomed Res Int. 2017;37(1):2–10. doi:10.1155/2017/2027594

5. Wan D, Krassioukov A V. Life-threatening outcomes associated with autonomic dysreflexia: A clinical review. J Spinal Cord Med. 2014. doi:10.1179/2045772313y.0000000098

6. Krassioukov A, Biering-Sorensen F, Donovan W, et al. International standards to document remaining autonomic function after spinal cord injury. Spinal Cord. 2012;35(4):201–210. doi:10.1038/sc.2008.121

7. Walter M, Knüpfer SC, Cragg JJ, et al. Prediction of autonomic dysreflexia during urodynamics: A prospective cohort study. BMC Med. 2018;16(1):53. doi:10.1186/s12916-018-1040-8

8. Sheel AW, Krassioukov A V., Inglis JT, Elliott SL. Autonomic dysreflexia during sperm retrieval in spinal cord injury: influence of lesion level and sildenafil citrate. J Appl Physiol. 2005;99(1):53–58. doi:10.1152/japplphysiol.00154.2005

9. Claydon VE, Elliott SL, Sheel AW, Krassioukov A. Cardiovascular responses to vibrostimulation for sperm retrieval in men with spinal cord injury. J Spinal Cord Med. 2006;29(3):207–216. doi:10.1080/10790268.2006.11753876

10. Brackett NL, Padron OF, Lynne CM. Semen Quality of Spinal Cord Injured Men is Better When Obtained by Vibratory Stimulation Versus Electroejaculation. J Urol. 1997;157(1):151–157. doi:10.1016/S0022-5347(01)65311-4

11. Ohl DA, SØnksen J, Menge AC, McCabe M, Keller LM. Electroejaculation versus vibratory stimulation in spinal cord injured men: Sperm quality and patient preference. J Urol. 1997;157(6):2147–2149. doi:10.1016/S0022-5347(01)64698-6

12. Davidson R, Elliott S, Krassioukov A. Cardiovascular Responses to Sexual Activity in Able-Bodied Individuals and Those Living with Spinal Cord Injury. J Neurotrauma. 2016;33(24):2161–2174. doi:10.1089/neu.2015.4143

13. Groen J, Pannek J, Castro Diaz D, et al. Summary of European Association of Urology (EAU) Guidelines on Neuro-Urology. Eur Urol. 2016;69(2):324–333. doi:10.1016/j.eururo.2015.07.071

14. Panicker JN, Fowler CJ, Kessler TM. Lower urinary tract dysfunction in the neurological patient: Clinical assessment and management. Lancet Neurol. 2015;14(7):720–732. doi:10.1016/S1474-4422(15)00070-8

15. Curt A, Nitsche B, Rodic B, Schurch B, Dietz V. Assessment of autonomic dysreflexia in patients with spinal cord injury. J Neurol Neurosurg Psychiatry. 1997;62(5):473–477. doi:10.1136/jnnp.62.5.473

16. Huang YH, Bih LI, Liao JM, Chen SL, Chou LW, Lin PH. Blood pressure and age associated with silent autonomic dysreflexia during urodynamic examinations in patients with spinal cord injury. Spinal Cord. 2013;51(5):401–405. doi:10.1038/sc.2012.155

17. Walter M, Knüpfer SC, Leitner L, et al. Autonomic dysreflexia and repeatability of cardiovascular changes during same session repeat urodynamic investigation in women with spinal cord injury. World J Urol. 2016. doi:10.1007/s00345-015-1589-1

18. Krassioukov A V., Weaver LC. Episodic hypertension due to autonomic dysreflexia in acute and chronic spinal cord-injured rats. Am J Physiol - Hear Circ Physiol. 1995;268(5):H2077–H2083. doi:10.1152/ajpheart.1995.268.5.h2077

19. Maiorov DN, Fehlings MG, Krassioukov A V. Relationship between severity of spinal cord injury and abnormalities in neurogenic cardiovascular control in conscious rats. J Neurotrauma. 1998;15(5):365–374. doi:10.1089/neu.1998.15.365

20. Scott MB, Morrow JW. Phenoxybenzamine in neurogenic bladder dysfunction after spinal cord injury. II. Autonomic dysreflexia. J Urol. 1978;119(4):483–484. doi:10.1016/S0022-5347(17)57524-2

21. Brown R, Stolzenhein G, Engel S, MacEfield VG. Cutaneous vasoconstriction as a measure of incipient autonomic dysreflexia during penile vibratory stimulation in spinal cord injury. Spinal Cord. 2009;47(7):538–544. doi:10.1038/sc.2008.158

22. Ekland MB, Krassioukov A V., McBride KE, Elliott SL. Incidence of autonomic dysreflexia and silent autonomic dysreflexia in men with spinal cord injury undergoing sperm retrieval: Implications for clinical practice. J Spinal Cord Med. 2008;31(1):33–39. doi:10.1080/10790268.2008.11753978

23. Solinsky R, Kirshblum SC, Burns SP. Exploring detailed characteristics of autonomic dysreflexia. J Spinal Cord Med. 2018;41(5):549–555. doi:10.1080/10790268.2017.1360434

24. NAFTCHI NE. Mechanism of Autonomic Dysreflexia. Ann N Y Acad Sci. 1990;579(1 A Decade of N):133–148. doi:10.1111/j.1749-6632.1990.tb48356.x

25. Hector SM, Biering-Sørensen T, Krassioukov A, Biering-Sørensen F. Cardiac arrhythmias associated with spinal cord injury. J Spinal Cord Med. 2013;36(6):591–599. doi:10.1179/2045772313Y.0000000114

26. Collins HL, Rodenbaugh DW, DiCarlo SE. Spinal cord injury alters cardiac electrophysiology and increases the susceptibility to ventricular arrhythmias. In: Progress in Brain Research. Vol 152. Elsevier; 2006:275–288. doi:10.1016/S0079-6123(05)52018-1

27. West CR, Bellantoni A, Krassioukov A V. Cardiovascular function in individuals with incomplete spinal cord injury: A systematic review. Top Spinal Cord Inj Rehabil. 2013;19(4):267–278. doi:10.1310/sci1904-267

28. Vírseda-Chamorro M, Salinas-Casado J, Gutiérrez-Martín P, de la Marta-García M, López-García-Moreno A, Esteban Fuertes M. Risk factors to develop autonomic dysreflexia during urodynamic examinations in patients with spinal cord injury. Neurourol Urodyn. 2017;36(1):171–175. doi:10.1002/nau.22906

29. Linsenmeyer TA, Campagnolo DI, Chou IH. Silent autonomic dysreflexia during voiding in men with spinal cord injuries. J Urol. 1996;155(2):519–522. doi:10.1016/S0022-5347(01)66438-3

