## supplementary methods for "Characterization of heart rate changes associated with autonomic dysreflexia during penile vibrostimulation and urodynamics"

This supplemental file provides additional information on the penile vibrostimulation (PVS) and urodynamic studies (UDS) including ethics, inclusion and exclusion criteria as well as associated assessments.

### PVS

**Ethics:**

- University of British Columbia Clinical Research Ethics Board (H13-02991)
- Vancouver Coastal Health Research Institute (V12-02991)

**Inclusion criteria:**

- Must be an English-speaking person of male sex
- Age must be >19 years to <55 years
- Participants must have an SCI with a complete or incomplete lesion above the 6$^{th}$ thoracic vertebral level
- The aforementioned SCI lesion must be >1 year's duration at the time of initial assessment
- A subjective experience of AD must have been previously elicited during self-stimulation or previous vibrostimulation
- Participants must have a proven ability to ejaculate vibratory stimulation in a clinical setting
- Must be able to provide written consent

**Exclusion criteria**:

- Participants who are not English-speaking
- People who are not of male sex.
- Participants who are <19 and >55 years of age
- Those with symptomatic cardiovascular disease including but not limited to: active coronary artery disease, angina, previous myocardial infarction, previous stroke or other high-risk cardiovascular events
- Those experiencing any of the following: unstable non-cardiovascular medical issues, a refractory psychiatric condition, substance use disorder, or alcohol use disorder, that will likely affect their ability to participant in, and complete the study
- Participants who have taken Tadalafil (Cialis) within 4 days of a scheduled study visit
- Participants who have taken Vardenafil (Staxyn or Levitra) or Sildenafil (Viagra) within 2 days of a scheduled study visit

**Study design:**

- prospective study

**Study location:**

- International Collaboration on Repair Discoveries, Blusson Spinal Cord Centre, 818 West 10th Avenue, Vancouver, British Columbia V5Z 1M9, Canada.

**Associated assessments:**

- The following assessments were modelled after the PVS protocol performed in Phillips et. al in 2018.[1]

*Injury assessment:*

- The International Standards for Neurological Classification of SCI (ISFNCSCI) examination was performed by a trained physician to confirm the neurological level of and completeness of injury in order to determine a grade on the American Spinal Injury Association Impairment Scale (AIS).[2]

*Penile Vibrostimulation for Sperm Retrieval:*

- On the day of the procedure, participants were instructed against any changes in routine
- The procedure was performed by a trained physician
- Penile vibrostimulation was applied to the glans penis using one or more handheld vibrators (Ferti Care, Multicept, Rungsted, Denmark) to induce ejaculation
- The frequency and amplitude of the vibration stimulus was targeting at 70-100 Hz and 1.0-3.5 mm, respectively
- Each vibration procedure was performed for a maximum of 3 minutes, with at least 30 seconds of elapsed time between stimulations
- Stimulated procedures were performed repeatedly until the participant ejaculated, or until at least five stimulations had elapsed

*Cardiovascular Monitoring and Assessment:*

- Baseline cardiovascular monitoring occurred in the ten minutes prior to, and throughout the Penile Vibrostimulation procedure, with data sampled at 1000 Hz using an analog-to-digital converter (powerlab/16SP ML 795; ADInstruments, Colorado Springs, CO) and transferred to data acquisition software (LabChart8; AD instruments, Colorado Springs, CO)
- Cardiovascular monitoring included the following: beat-to-beat blood pressure monitoring using non-invasive finger plethysmography (Finometer PRO; Finapres Medicine Systems, Amsterdam, Netherlands), and electrocardiogram (ML132; ADInstruments, Colorado Springs, CO)
- Participants were also asked to comment on: whether their AD symptoms were present, the severity of these symptoms, and whether their symptoms were comparable to personal situations
- This information was added in real-time during the procedure to the data acquisition software recordings

**UDS**

**Ethics:**

- University of British Columbia Clinical Research Ethics Board (H15-02364)
- Vancouver Coastal Health Research Institute (V15-02364)
- Health Canada (205857)
- Clinicaltrials.gov registration: NCT02676154

**Inclusion criteria:**

- Must be >18yrs and <60 years of age
- Must be English speaking or have access to an appropriate interpreter
- Participants can be of male or female sex
- Participants must have an SCI with a complete or incomplete lesion at or above the 6th thoracic vertebral level
- The aforementioned SCI lesion must be >1 year's duration at the time of initial assessment
- Must have documented presence of AD and NDO during clinical UDS assessments
- Participants must have hand function that is sufficient to perform CIC or, a designated caregiver to provide regular CIC
- Participants must comply with the required documentation of bladder and bowel function in the 2 weeks prior to their initial baseline visit
- Must be willing and able to comply with all scheduled clinic visits and study-related procedures
- Must be able to understand and complete study-related questionnaires
- People who are of female sex and childbearing age must not be: intending to become pregnant, currently pregnant, or breast-feeding
- Sexually active participants (female sex of childbearing age or male sex with partners who are female sex of childbearing age) must agree to use effective contraception during the period of the trial and for at least 28 days after completion of treatment
- Must provide informed consent prior to participation

**Exclusion criteria:**

- Female sex participants who are: planning to become pregnant, currently pregnant, or breastfeeding
- Those with the following medical conditions which preclude participation, including:
    - Chronic kidney disease (GFR <60 ml/min) and hepatic disease (abnormal liver enzyme and liver function tests)
    - active urinary tract infection with risk of progression to pyelonephritis
    - poorly-controlled Type 1 or 2 diabetes mellitus
    - active pressure sores, active heterotopic ossification, or recent changes to anti-depressant prescriptions
- Otherwise, participation was precluded in anyone with a severe acute medical issue that was determined to affect the patient's participation in the study

- Must not be hypersensitive to Tolterodine (available as Detrol, Detrol LA), soy, peanuts or lactose
- Must not have had recent treatment of the detrusor muscle with botulinumtoxinA (Botox) within 9 months of the first baseline visit
- No recent treatment with other anticholinergic medications within 3 weeks of the baseline visit
- No use of any medication or treatment the precludes safe participation in the study, including ketoconazole, itraconazole, miconazole, clarithromycin, or rifampicin
- The participant must not be a member of the investigational team or is an immediate family member of the investigational team

**Study design:**

- phase II, open-label exploratory, non-blinded, non-randomized, single-centre study

**Study location:**

- International Collaboration on Repair Discoveries, Blusson Spinal Cord Centre, 818 West 10th Avenue, Vancouver, British Columbia V5Z 1M9, Canada.

**Associated Assessments:**

*Injury assessment:*

- The ISFNCSCI examination was performed by a trained physician to confirm the neurological level of and completeness of injury in order to determine a grade on the AIS[2]

*UDS:*

- All methods, definitions and units regarding UDS are in accordance with the standards recommended by the International Continence Society (ICS)[3]
- Urodynamics will be performed according to good urodynamic practices recommended by the ICS to detect/confirm NDO[4]
- Confirmation of AD during UDS: an attending physician will confirm episodes of AD, which is defined according to the ISAFSCI[5] as an increase in SBP ≥20 mmHg from baseline
- Patients were also asked to empty their bladder and bowels prior to the study, in accordance with the International Continence Society's Good Urodynamic Practice guidelines[4]
- The urodynamic system (Aquarius TT, Laborie Model 94-R03-BT, Montreal, QC, Canada) used to perform the assessment was also used in accordance with these guidelines

*Cardiovascular monitoring during UDS:*

- Concurrent to UDS, we continuously recorded beat-by-beat blood pressure, via finger photoplethysmography (Finometer PRO, Finapres Medical Systems, Amsterdam, Netherlands) corrected to brachial pressure (CARESCAPE V100, GE Healthcare, Milwaukee, WI, USA), and one-lead electrocardiogram (eML 132; ADInstruments, Colorado Springs, CO, USA) for heart rate in order to detect autonomic dysreflexia[6]

**Data recording extraction**

For both studies, HR, SBP, and DBP were directly recorded during the entire protocol using beat-to-beat plethysmography and MAP was calculated (MAP = [SBP + 2 $\times$ DBP]/3). Baseline data for both PVS and UDS was acquired from the minute prior to each procedure, and additionally in the case of the PVS study, between stimulations for 30 seconds to re-establish a new baseline each time. Analysis of the live recordings began with a 3-beat average for SBP, DBP and MAP. HR data was calculated from the same heartbeats used for BP extraction. Data was collected for each point of interest during the study, including recorded AD symptoms, lack of symptoms, other adjustments made, and points specific to each type of procedure (i.e. the point of ejaculation in PVS, and catheter adjustments made in UDS).

**REFERENCES**


1. Phillips AA, Matin N, Jia M, et al. Transient Hypertension after Spinal Cord Injury Leads to Cerebrovascular Endothelial Dysfunction and Fibrosis. *J Neurotrauma*. 2018;35(3):573-581. doi:10.1089/neu.2017.5188

2. Kirshblum SC, Burns SP, Biering-Sorensen F, et al. International standards for neurological classification of spinal cord injury. *J Spinal Cord Med*. 2011;34(6):535-546. doi:10.1179/204577211X13207446293695

3. Abrams P, Cardozo L, Fall M, et al. The standardisation of terminology of lower urinary tract function: Report from the standardisation sub-committee of the international continence society. *Am J Obstet Gynecol*. 2002. doi:10.1067/mob.2002.125704

4. Schäfer W, Abrams P, Liao L, et al. Good urodynamic practices: Uroflowmetry, filling cystometry, and pressure-flow studies**. *Neurourol Urodyn*. 2002;21(3):261-274. doi:10.1002/nau.10066

5. Krassioukov A, Biering-Sorensen F, Donovan W, et al. International standards to document remaining autonomic function after spinal cord injury. *Spinal Cord*. 2012;35(4):201-210. doi:10.1038/sc.2008.121

6. Walter M, Lee AHX, Kavanagh A, et al. Epidural Spinal Cord Stimulation Acutely Modulates Lower Urinary Tract and Bowel Function Following Spinal Cord Injury: A Case Report. *Front Physiol*. 2018;9:1816. doi:10.3389/fphys.2018.01816
